# Investigation of locomotive syndrome improvement by total hip arthroplasty in patients with hip osteoarthritis: a before-after comparative study focusing on 25-question geriatric locomotive function scale

**DOI:** 10.1101/2024.11.27.24318045

**Authors:** Shigeaki Miyazaki, Kurumi Tsuruta, Saori Yoshinaga, Yoshinori Fujii, Amy Hombu, Taro Funamoto, Takero Sakamoto, Takuya Tajima, Yoshihiro Nakamura, Hideki Arakawa, Jun Nakatake, Etsuo Chosa

## Abstract

**Background:** The 25-Question Geriatric Locomotive Function Scale (GLFS-25) is one of the tests used to assess the risk of locomotive syndrome (LS). It is a comprehensive tool to measure LS improvement after total hip arthroplasty (THA) and provides beneficial information for rehabilitation after THA. This study aims to investigate the effectiveness of LS improvement three months after THA in patients who underwent THA for hip osteoarthritis (OA) by using the total clinical decision limits (CDL) 3-stage scale to identify specific trends in LS improvement based on the GLFS-25.

**Methods:** The participants of this study were 273 patients who underwent primary THA for hip OA. LS was evaluated using the stand-up test, two-step test, and GLFS-25 before receiving THA and three months after THA.

**Results:** The total CDL was improved by THA in 132 patients (improvement rate: 48.4%). There were 15 questions out of 25 in the GLFS-25 showed improvements in more than 60% of the patients. The question Q13, “To what extent has it been difficult to walk briskly?” revealed the highest improvement rate, followed by Q12, “To what extent has it been difficult to go up and down stairs?”, and Q3, “Did you have any pain (including numbness) in your lower limbs?”.

**Conclusions:** At three months after THA, approximately half of the patients showed improvement in LS. Furthermore, improvement in locomotion movements was identified as the most significant area of progress among motor functions.

## Introduction

The aging population is a significant issue around the world. The number of people aged 60 and over reached 1 billion in 2019. This number is expected to reach 1.4 billion by 2030 and 2.1 billion by 2050. This increase will continue to accelerate in the coming decades, especially in developing countries [1]. The number of elderly people requiring nursing care is expanding due to motor dysfunction [2]. Ensuring the elderly can maintain a good quality of life (QOL) can benefit both individuals and society.

Locomotive syndrome (LS) was introduced by the Japanese Orthopedic Association (JOA) in 2007 to evaluate motor functions due to locomotive organ impairment [3]. LS leads to the risk of requiring future long-term health care [4–6]. In 2013, JOA developed the LS risk tests consisting of a stand-up test, two-step test, and 25-Question Geriatric Locomotive Function Scale (GLFS-25) to assess LS [7]. Additionally, in 2015, JOA implemented clinical decision limits (CDL) as criteria for determining the risks of LS [8]. The CDL classifies the LS into stage 1 or stage 2 [7]; in 2020, stage 3 was added, making CDL a three-stage evaluation instrument [9,10].

Total hip arthroplasty (THA) is a surgical treatment to regain hip joint function, relieve pain, and improve the limitations of activities of daily living (ADL) in patients with hip osteoarthritis (OA). THA has shown effectiveness in enhancing QOL, including walking function [11], sports activities [12], and cardiopulmonary function [13], which leads to early reintegration into society. Larsen et al. [14] reported that early perioperative care and rehabilitation can significantly shorten hospital stay and improve QOL three months after THA. Furthermore, Miyazaki et al. [9] reported that the improvement rate in total CDL three months after THA was 46.7%, indicating that patients could recover to independent walking three months after THA. The demonstration of the effectiveness in improving LS based on the short-term results due to THA is clinically significant.

The GLFS-25, one of the LS risk tests, was developed as a screening instrument for elderly patients with motor dysfunctions [15, 16]. A comprehensive consideration of LS improvement after THA using the GLFS-25 provides beneficial information for implementing after THA rehabilitation. Regarding the effect of THA on improving the GLFS-25, previous studies reported that 90% of patients with severe hip joint disorders required nursing care before THA. This percentage dropped to 30% three months after THA [17]. Unfortunately, up to date, no short-term results at three months after THA in hip OA patients have been reported concerning the effect of LS improvement using the total CDL stages 1 to 3. Furthermore, there is no study to examine the trend or degree of progress for each question in the GLFS-25. This study aims to investigate the effectiveness of LS improvement at three months after THA in patients who underwent THA for hip OA by using the total CDL 3-stage scale to identify specific trends in LS improvement based on the GLFS-25.

## Materials and Methods

### Study design and ethical statement

This before-after comparative study was approved by the Research Ethics Committee of the University of Miyazaki, School of Medicine (Approval No. O-0783). It was carried out in compliance with the Ethical Guidelines for Medical and Biological Research Involving Human Subjects at the Department of Rehabilitation Medicine, University of Miyazaki Affiliated Hospital. The participants of this study were patients who underwent primary THA for hip OA between October 2018 and May 2024. Approval for this study was obtained from the Research Ethics Committee in September 2020. Therefore, data collected before obtaining approval from the Research Ethics Committee were categorized as a retrospective study, while data collected after obtaining approval were categorized as a prospective study. All data used in the retrospective study were fully anonymized prior to access. As it was not feasible to contact participants in the retrospective study and because the information was initially collected for clinical purposes, the opt-out method was applied in accordance with ethical guidelines. Information regarding the conduct of the research including the objectives was disclosed and the research participants were provided an opportunity to refuse inclusion in the research.

### Patient selection

The participants of this study were patients who underwent primary THA for hip OA. The target patients agreed to participate in both before THA and three months after THA evaluations. Patients with femoral head necrosis, trauma, rheumatoid arthritis, infection, and patients with incomplete outcome measure data sets were excluded from the study. As a result of careful selection and rigorous screening, 273 patients were included in this study (Fig 1). The breakdown of the total CDL stages was: 0 patients of stage 0, 1 patients of stage 1, 27 patients of stage 2, and 245 patients of stage 3. The cohort in this study included patients with polyarticular disease and those who have undergone arthroplasty in other joints.

**Fig 1.**
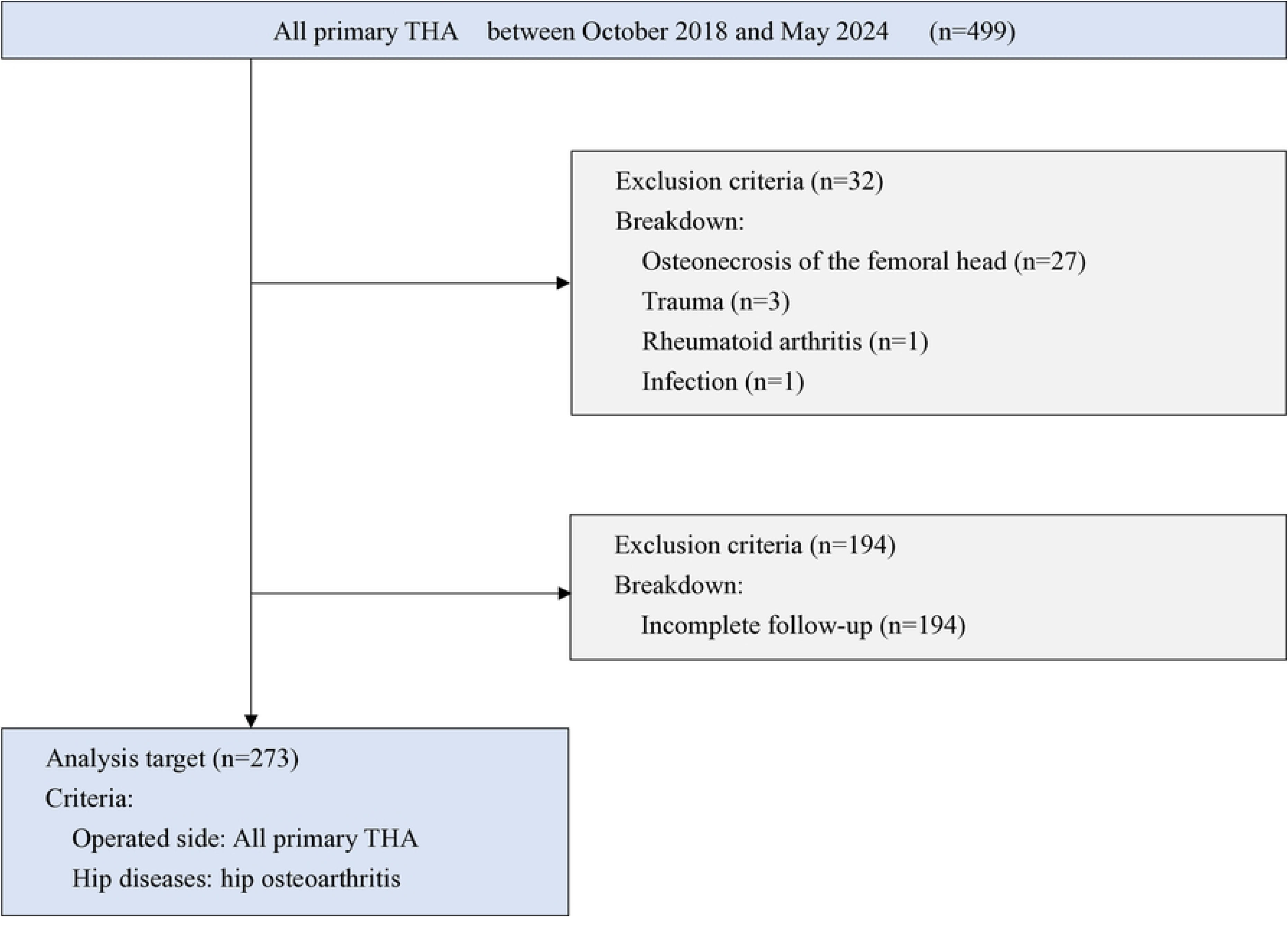
Flowchart of the included patients. The participants of this study were patients who underwent primary THA for hip OA between October 2018 and May 2024. Patients with femoral head necrosis, trauma, rheumatoid arthritis, infection, and patients with incomplete outcome measure data sets were excluded from the study.

All cases in this study underwent THA using the anterior minimally invasive surgery (AMIS) approach or transgluteal approach performed by experienced orthopedic surgeons at the hospital of the authors’ affiliated institution. Rehabilitation after THA started the day after THA for three months. Patients received one-on-one rehabilitation twice a day for 60 to 80 minutes during hospitalization. Outpatients received one-on-one rehabilitation for 20 to 40 minutes two to three times a week.

### Outcome measures

The three LS risk tests proposed by JOA, the stand-up test, two-step test, GLFS-25, and total CDL stage, were adopted as outcome measures in this before-after comparative study [7]. For evaluation, the CDL and total CDL results of each test were classified into stages 0 to 3. All measures were performed both before and three months after THA. Data were collected as previously described in Miyazaki et al. [10].

1. For the stand-up test, following the JOA guidelines, four different stool heights were used: 40 cm, 30 cm, 20 cm, and 10 cm. First, a patient was asked to stand up from the sitting position starting at the highest stool of 40 cm. After being able to stand up with both legs, the next step was to stand up with one leg. After that, the test continues using stools at different heights. The evaluation employed a 9-point scale [7]: 0 points (unable to stand); 1, 2, 3, and 4 points (stand on both legs from heights of 40 cm, 30 cm, 20 cm, and 10 cm, respectively); 5, 6, 7, and 8 points (stand on one leg from heights of 40 cm, 30 cm, 20 cm, and 10 cm, respectively). Scores <2, <3, and <5 were classified as CDL stages 3, 2, and 1, respectively. The instructions to quantify motion were given. (1) Fold arms in front of the chest at a sitting position. (2) Place feet shoulder-width apart. (3) Position lower legs at an angle of approximately 70° to the floor. (4) Stand up without gaining momentum. (5) Maintain the standing posture for 3 seconds.
2. The two-step test is used to evaluate stride length. A patient was asked to take two steps with the longest possible stride, and the stride length of these two steps was measured. The test score was calculated by dividing the stride length of these two steps by the patient’s height. Scores <0.9, ≥0.9 to <1.1, and ≥1.1 to <1.3 were classified into CDL stages 3, 2, and 1, respectively. The instructions to quantify motion were given. (1) Align toes at the starting line with a stationary standing position. (2) Take two of the longest possible steps forward, then align toes together in a stationary standing position. (3) If the subject loses balance, start over. (4) Measure the stride length of the two steps. (5) Perform two times and adopt the better score.
3. The GLFS-25 [15] is a self-administered assessment of 25 questions. This test consists of four questions measuring physical pain, 16 questions measuring ADL, three questions measuring social functions, and two questions measuring mental health in the past month before the test. These 25 questions are rated on a 5-point scale ranging from no functional impairment (0 points) to severe functional impairment (4 points), and a total score is calculated (minimum 0 points, maximum 100 points). Scores ≥24, ≥16 to <24, and ≥7 to <16 were classified into CDL stages 3, 2, and 1, respectively.
4. The total CDL was determined based on the results of the stand-up test, two-step test, and GLFS-25. As a result, the most advanced stage of mobility impairment can be determined based on the CDL stages.

### Statistical Analysis

The following statistical analyses were performed.

1. Evaluation of LS improvement: The Wilcoxon signed-rank test was used to compare the distribution of the CDL stages for the three LS risk tests and the total CDL stages.
2. Changes in each of the LS test scores and each of the GLFS-25 questions before THA and three months after THA were examined using a Paired t-test.

This is an observational study with a longitudinal design. At the time of applying for ethical approval, we could not predict how many cases we would have over the 6-year period. As a result, we had 273 participants eligible for this study, all of whom underwent primary THA at the hospital during the study period, excluding those who did not meet the patient selection criteria. All statistical analyses were performed using IBM SPSS 27.0 (IBM Corp., Released 2020. Armonk, NY, USA). The statistical significance was set at *p* < .05.

## Results

Table 1 shows the age, sex, height, weight, and BMI. The participants were 273 patients comprising 52 males and 221 females, and the mean age was 67.4 years (standard deviation SD: 9.3 years). The mean BMI was 24.4 kg/m^2^ (standard deviation SD: 3.8 kg/m^2^).

**Table 1.** Characteristics of the patients.

| Table 1. Characteristics of the patients. |  |
| --- | --- |
| Characteristic | Total (n = 273) |
| Age (years) | 67.4 ± 9.3 |
| Sex: male/female | 52 / 221 |
| Height (cm) | 154.3 ± 7.6 |
| Weight (kg) | 58.4 ± 11.0 |
| Body mass index (BMI) (kg/m<sup>2</sup>) | 24.4 ± 3.8 |
| BMI, body mass index. |  |
| Age, height, weight, and BMI values are means ± standard deviation. |  |

Table 2 shows the participants’ medical information. Among the participants in this study, 53 patients (19.4%) had primary hip OA, and 220 (80.6%) patients had secondary hip OA. Regarding THA, 217 (79.5%) patients were unilateral, and 56 (20.5%) patients were bilateral. Regarding the surgical procedure, 156 (57.1%) patients had the AMIS approach, and 117 (42.9%) patients had the transgluteal approach. Regarding the stage on the non-operated side, in the order of stages 0 to 4, the details were 39 (14.3%), 52 (19.0%), 39 (14.3%), 35 (12.8%), and 45 (16.5%) patients, respectively, and 63 (23.1%) were after THA. On the operated side, in the order of stages 0 to 4, the details were 0 (0%), 0 (0%), 0 (0%), 33 (12.1%), and 240 (87.9%) patients, respectively.

**Table 2.**
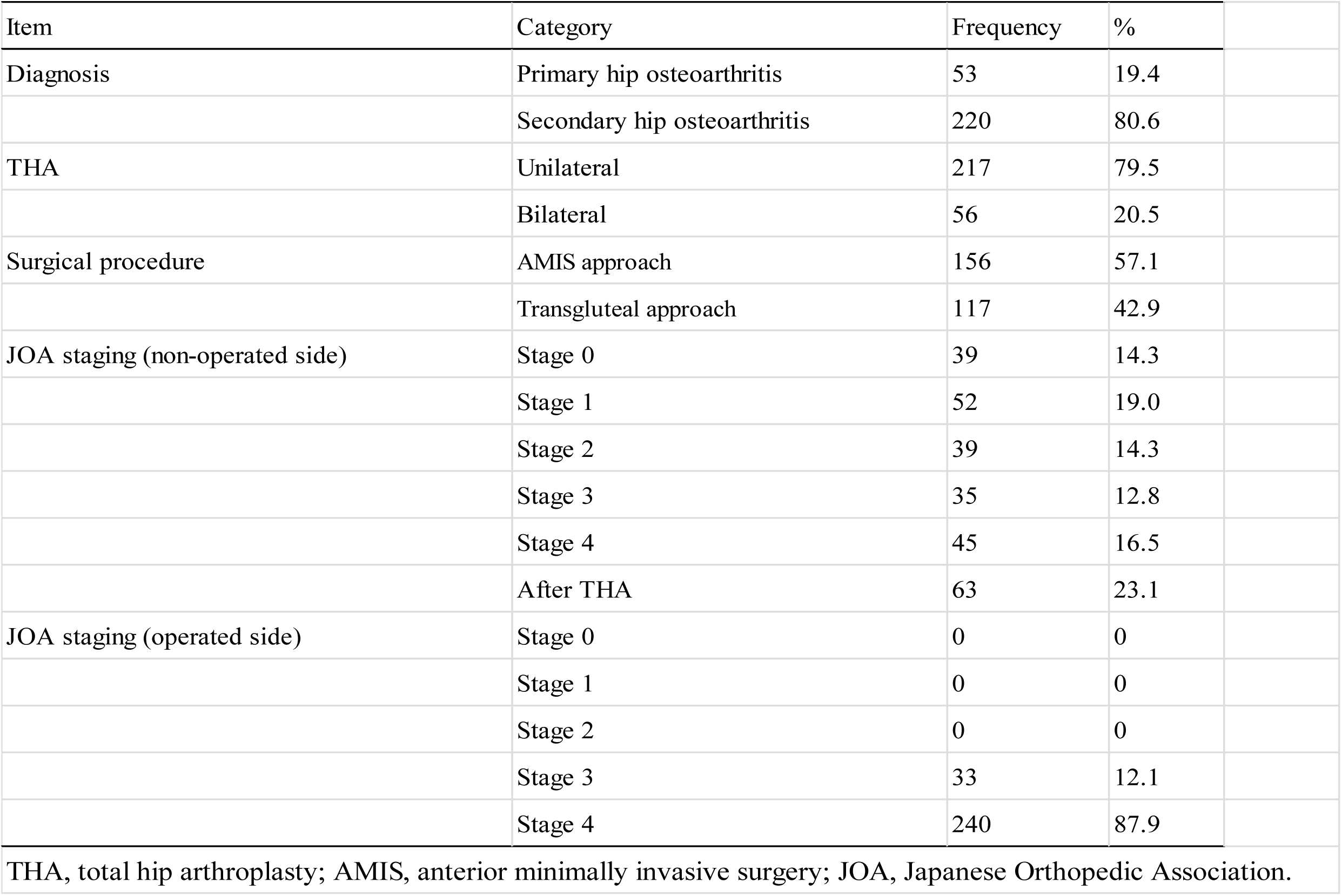
Patients’ medical information.

| Table 2. Patients' medical information. |  |  | n = 273 |
| --- | --- | --- | --- |
| Item | Category | Frequency | % |
| Diagnosis | Primary hip osteoarthritis | 53 | 19.4 |
|  | Secondary hip osteoarthritis | 220 | 80.6 |
| THA | Unilateral | 217 | 79.5 |
|  | Bilateral | 56 | 20.5 |
| Surgical procedure | AMIS approach | 156 | 57.1 |
|  | Transgluteal approach | 117 | 42.9 |
| JOA staging (non-operated side) | Stage 0 | 39 | 14.3 |
|  | Stage 1 | 52 | 19.0 |
|  | Stage 2 | 39 | 14.3 |
|  | Stage 3 | 35 | 12.8 |
|  | Stage 4 | 45 | 16.5 |
|  | After THA | 63 | 23.1 |
| JOA staging (operated side) | Stage 0 | 0 | 0 |
|  | Stage 1 | 0 | 0 |
|  | Stage 2 | 0 | 0 |
|  | Stage 3 | 33 | 12.1 |
|  | Stage 4 | 240 | 87.9 |

THA, total hip arthroplasty; AMIS, anterior minimally invasive surgery; JOA, Japanese Orthopedic Association.

Table 3 shows the distributions of the CDL stages for the three LS tests and the total CDL stages for all participants. There was a significant change in the percentage of participants in each CDL stage from before THA to three months after THA (*p* < .001). At three months, the percentage of participants with total CDL stage 3 decreased, while the percentage of participants with total CDL stages 2, 1, and 0 increased. The same trend was detected in the distribution for each of the three tests in LS.

**Table 3.** Distribution of the CDL stage for the three LS tests and the total CDL stages (n = 273).

|  |  |  |  |  |  |  |  |  |  |  |
| --- | --- | --- | --- | --- | --- | --- | --- | --- | --- | --- |
|  |  | Stage 0 |  | Stage 1 |  | Stage 2 |  | Stage 3 |  | Wilcoxon signed-rank test |
|  |  | N | % | N | % | N | % | N | % | *p*-value |
| Stand-up test | Before THA | 7 | 2.6 | 84 | 30.8 | 82 | 30.0 | 100 | 36.6 | < 0.001*** |
|  | 3 months after THA | 16 | 5.9 | 98 | 35.9 | 92 | 33.7 | 67 | 24.5 |  |
| Two-step test | Before THA | 3 | 1.1 | 36 | 13.2 | 101 | 37.0 | 133 | 48.7 | < 0.001*** |
|  | 3 months after THA | 14 | 5.1 | 80 | 29.3 | 115 | 42.1 | 64 | 23.4 |  |
| GLFS-25 | Before THA | 0 | 0.0 | 5 | 1.8 | 34 | 12.5 | 234 | 85.7 | < 0.001*** |
|  | 3 months after THA | 32 | 11.7 | 91 | 33.3 | 65 | 23.8 | 85 | 31.1 |  |
| Total CDL stage | Before THA | 0 | 0.0 | 1 | 0.4 | 27 | 9.9 | 245 | 89.7 | < 0.001*** |
|  | 3 months after THA | 2 | 0.7 | 51 | 18.7 | 93 | 34.1 | 127 | 46.5 |  |
| CDL, clinical decision limits; LS, locomotive syndrome; THA, total hip arthroplasty; GLFS-25, 25-Question Geriatric Locomotive Function Scale. |  |  |  |  |  |  |  |  |  |  |
| Significantly different: *** *p* < .001 |  |  |  |  |  |  |  |  |  |  |

Table 4 shows the changes observed in total CDL due to THA. THA improved the total CDL in 132 patients with an improvement rate of 48.4%. Before THA, 27 patients were in LS stage 2, of which seven (25.9%) improved to LS stage 1, and one (3.7%) to LS stage 0 after THA. Similarly, among 246 patients who were LS stage 3 before THA, 80 (32.5%) improved to LS stage 2, 43 (17.5%) to LS stage 1, and one (0.4%) to LS stage 0.

**Table 4.** Changes in the total CDL stage by THA.

| **Table 4. Changes in the total CDL stage by THA.** |  |  |  |  |  |  |  |  |  |  |
| --- | --- | --- | --- | --- | --- | --- | --- | --- | --- | --- |
| 3 months<br>after THA<br>Before THA | Stage 0 |  | Stage 1 |  | Stage 2 |  | Stage 3 |  | Total |  |
|  | N | % | N | % | N | % | N | % | N | % |
| Stage 2 | **1** | 3.7 | **7** | 25.9 | 13 | 48.2 | 6 | 22.2 | 27 | 100 |
| Stage 3 | **1** | 0.4 | **43** | 17.5 | **80** | 32.5 | 122 | 49.6 | 246 | 100 |
| Total | 2 | 0.7 | 50 | 18.3 | 93 | 34.1 | 128 | 46.9 | 273 | 100 |
| Highlighted numbers indicate improvements (132 patients, 48.4% improvement rate) |  |  |  |  |  |  |  |  |  |  |
| CDL, clinical decision limits; THA, total hip arthroplasty. |  |  |  |  |  |  |  |  |  |  |

Table 5 shows the significant differences (*p* < .001) observed in the scores for all cases in the three LS risk tests (stand-up test, two-step test, and GLFS-25) between before THA and three months after THA. The scores of the stand-up test and the two-step test were significantly higher at three months after THA. The GLFS-25 score was significantly lower at three months after THA.

**Table 5.** Changes in the three LS test scores and each functional parameter from before to 3 months after THA (n = 273).

|  |  |  |  |  |  |
| --- | --- | --- | --- | --- | --- |
| | Before THA<br>(mean ± SD) | 3 months after THA<br>(mean ± SD) | Paired *t*-test<br>*p*-value | | |
| Stand-up test | 2.06 ± 1.21 | 2.36 ± 1.17 | < 0.001*** | | |
| Two-step test | 0.88 ± 0.22 | 1.01 ± 0.20 | < 0.001*** | | |
| GLFS-25 | 40.9 ± 16.0 | 19.3 ± 12.4 | < 0.001*** | | |
| LS, locomotive syndrome; THA, total hip arthroplasty; SD, standard deviation; |  |  |  |  |  |
| GLFS-25, 25-Question Geriatric Locomotive Function Scale. |  |  |  |  |  |
| Significantly different: *** *p* < .001 |  |  |  |  |  |

The differences in the GLFS-25 scores before THA and three months after THA are shown using a scatter plot (Fig 2). There is a negative correlation between the GLFS-25 score before THA and the change in the GLFS-25 score (three months after THA minus before THA), and the change in the GLFS-25 score increased in proportion to the GLFS-25 score before THA. As a result of regression analysis, the regression coefficient was – 0.723 and the coefficient of determination (R^2^) was 0.498 (*p* < .001).

**Figure 2.**
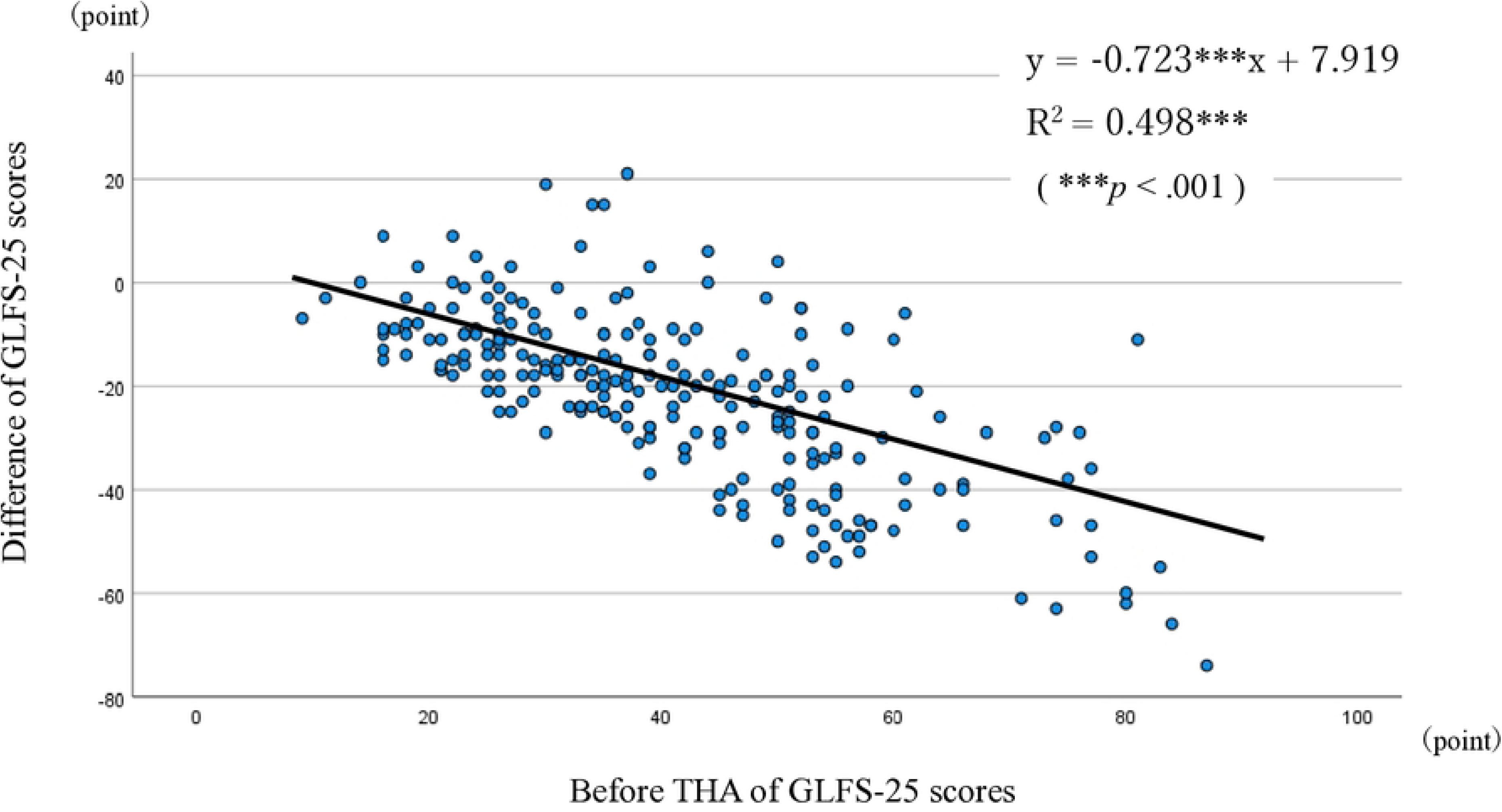
Scatter plot graph of GLFS-25 score. The scatter plot shows the correlation between the GLFS-25 score before THA and the change in the GLFS-25 score (three months after THA minus before THA).

Table 6 shows the average score difference between before and three months after THA for each GLFS-25 question and the percentage of patients who improved in three months after THA compared to the before THA evaluation. For all questions, there was a significant difference between the scores before THA and three months after THA; in other words, the scores three months after THA were lower than before THA. Furthermore, the percentage of patients who improved was over 60% for 15 out of the 25 questions, and the question with the highest percentage of patients who improved was Q13 (80.2%). This was followed by Q12 (76.9%), Q3 (76.2%), Q4 (74.7%), Q7 (72.5%), and Q21 (70.7%). On the other hand, the question with the lowest improvement rate was Q8 (24.5%).

**Table 6.** Changes in GLFS-25 and the improvement rate.

| Items | Before THA<br>(mean ± SD) | 3 months after THA<br>(mean ± SD) | Difference<br>(mean ± SE) | Improvement rate (%) |
| --- | --- | --- | --- | --- |
| Q1. Did you have any pain (including numbness) in your neck or upper limbs? | 0.78 ± 0.92 | 0.66 ± 0.82 | 0.12 ± 0.06* | 30.8 |
| Q2. Did you have any pain in your back, lower back or buttocks? | 1.32 ± 1.08 | 0.90 ± 0.92 | 0.42 ± 0.07 *** | 46.2 |
| Q3. Did you have any pain (including numbness) in your lower limbs? | 2.23 ± 1.06 | 0.89 ± 0.83 | 1.33 ± 0.08 *** | 76.2 |
| Q4. To what extent has it been painful to move your body in daily life? | 2.09 ± 0.92 | 0.82 ± 0.78 | 1.27 ± 0.07 *** | 74.7 |
| Q5. To what extent has it been difficult to get up from a bed or lie down? | 1.29 ± 0.99 | 0.43 ± 0.65 | 0.85 ± 0.07 *** | 60.4 |
| Q6. To what extent has it been difficult to stand up from a chair? | 1.35 ± 0.99 | 0.44 ± 0.62 | 0.91 ± 0.06 *** | 60.4 |
| Q7. To what extent has it been difficult to walk inside the house? | 1.36 ± 0.95 | 0.34 ± 0.58 | 1.03 ± 0.06 *** | 72.5 |
| Q8. To what extent has it been difficult to put on and take off shirts? | 0.41 ± 0.77 | 0.14 ± 0.39 | 0.27 ± 0.05 *** | 24.5 |
| Q9. To what extent has it been difficult to put on and take off trousers and pants? | 1.80 ± 0.96 | 0.76 ± 0.65 | 1.04 ± 0.06 *** | 68.5 |
| Q10. To what extent has it been difficult to use the toilet? | 0.87 ± 0.90 | 0.21 ± 0.44 | 0.66 ± 0.06 *** | 50.5 |
| Q11. To what extent has it been difficult to wash your body in the bath? | 1.11 ± 0.97 | 0.47 ± 0.62 | 0.65 ± 0.06 *** | 49.1 |
| Q12. To what extent has it been difficult to go up and down stairs? | 2.37 ± 0.99 | 1.06 ± 0.84 | 1.31 ± 0.07 *** | 76.9 |
| Q13. To what extent has it been difficult to walk briskly? | 2.62 ± 1.02 | 1.16 ± 0.89 | 1.46 ± 0.07 *** | 80.2 |
| Q14. To what extent has it been difficult to keep yourself neat? | 1.01 ± 0.94 | 0.34 ± 0.57 | 0.67 ± 0.06 *** | 50.9 |
| Q15. How far can you keep walking without rest? | 2.67 ± 1.12 | 1.52 ± 1.16 | 1.14 ± 0.08 *** | 69.2 |
| Q16. To what extent has it been difficult to go out to visit neighbors? | 1.44 ± 1.09 | 0.40 ± 0.66 | 1.04 ± 0.07 *** | 67.8 |
| Q17. To what extent has it been difficult to carry objects weighing approximately 2 kilograms? | 1.73 ± 1.23 | 0.82 ± 0.93 | 0.90 ± 0.08 *** | 60.8 |
| Q18. To what extent has it been difficult to go out using public transportation? | 2.00 ± 1.26 | 0.86 ± 0.98 | 1.14 ± 0.08 *** | 66.7 |
| Q19. To what extent have simple tasks and housework been difficult? | 1.13 ± 0.95 | 0.39 ± 0.63 | 0.74 ± 0.06 *** | 57.9 |
| Q20. To what extent have load-bearing tasks and housework been difficult? | 1.81 ± 1.17 | 0.92 ± 0.89 | 0.89 ± 0.07 *** | 60.1 |
| Q21. To what extent has it been difficult to perform sports activity? | 2.93 ± 1.03 | 1.70 ± 1.16 | 1.23 ± 0.08 *** | 70.7 |
| Q22. Have you been restricted from meeting your friends? | 1.36 ± 1.32 | 1.00 ± 1.18 | 0.36 ± 0.08 *** | 38.1 |
| Q23. Have you been restricted from joining social activities ? | 2.29 ± 1.46 | 1.66 ± 1.48 | 0.63 ± 0.10*** | 48.0 |
| Q24. Have you ever felt anxious about falls in your house? | 1.30 ± 1.05 | 0.76 ± 0.77 | 0.54 ± 0.07 *** | 44.3 |
| Q25. Have you ever felt anxious about being unable to walk in the future? | 1.68 ± 1.16 | 0.64 ± 0.76 | 1.04 ± 0.08 *** | 64.8 |
| Improvement rate is the comparison of before THA and 3 months after THA |  |  |  |  |
| GLFS-25, 25-Question Geriatric Locomotive Function Scale; THA, total hip arthroplasty; SD, standard deviation; SE, standard error. |  |  |  |  |
| Paired *t* -test, Significantly different: * *p* < .05, *** *p* < .001 |  |  |  |  |

## Discussion

This study evaluated patients with hip OA who underwent primary THA for each CDL stage and total CDL stage of LS risk tests before and three months after THA to assess the effectiveness of THA. In addition, the specific trends of LS improvement in motor function based on the GLFS-25 were examined. To the best of our knowledge, there have been no short-term evaluations at three months after THA in hip OA patients, including evaluation by total CDL stage 1 to 3 or evaluation of LS improvement after THA based on the GLFS-25. Therefore, this study is the first to clarify the therapeutic effects of THA on LS and the motor function questions related to LS improvement from the GLFS-25, focusing on three months after THA. The most important findings from this study were that total CDL improved in 48.4% of patients three months after THA; 15 out of 25 (60%) GLFS-25 questions revealed an improvement rate of over 60% in patients.

THA has been a well-received surgery that rebuilds hip joint function and improves pain and restrictions in ADL due to locomotive organ impairment. Previous studies reported implant survival rates exceeding 20 years after surgery [18, 19]. Consequently, it is rather crucial to clarify the improvement rate of total CDL after THA. In a cohort study examining the effects of THA in hip OA patients, the improvement rate in total CDL stage 1 and stage 2 evaluation was 57% at six months and 65% at one year after THA [20]. In a 2-year post-THA cohort study, the improvement rate in total CDL stage 1 and stage 2 evaluation was 56.8% [21]. In addition, a cohort study examining the short-term effects of THA reported that the improvement rate when evaluating total CDL stage 3 alone was 46.7% at three months after THA [9]. Our study is the first to evaluate total CDL in all stages, 1 to 3. Although it is not realistic to conduct a simple comparison with previous reports due to the time frame differences, our total CDL improvement rate three months after THA was 48.4% which was in line with other studies. Regarding the improvement change by total CDL stage, the improvement rate from stage 3 to stage 2, 1, or 0 was 50.4%, and some patients did show an improvement from stage 3 to 0. The improvement rate from stage 2 to 1 or 0 was 29.6%. This is to say that THA due to hip OA can improve LS within a short period of time after surgery.

In the present study, we investigated the specific trends of improvement by THA using the GLFS-25. The results showed that there was a negative correlation between the GLFS-25 score pre-THA and the change in the GLFS-25 score (three months after THA minus before THA). The changes seen in the scores toward the negative direction (-) indicate improvement in most patients. Furthermore, the higher the GLFS-25 score before THA and the higher the difficulty in ADL, the greater the improvement in score. Regarding the improvement effect of THA on the GLFS-25, Maezawa et al. [17] reported that the GLFS-25 improved from 55.4 points before THA to 19.1 points three months after THA. Our present study showed that the CDL stage in the GLFS-25 significantly decreased from 234 (85.7%) patients with CDL stage 3 before THA to 85 (31.1%) patients three months after THA. A significant improvement in the GLFS-25 score was also observed by 21.6 points (40.9 points before THA minus 19.3 points three months after THA).

The GLFS-25 assesses the following four categories: pain-related (Q1 to 4), ADL-related (Q5 to 20), social function-related (Q21 to 23), and mental health-related (Q24, 25) [15]. Both Q3 and Q4 correspond to hip joint pain-related. The score for both Q3 and Q4 was improved from two points (moderate pain) or higher before THA to less than one point (little pain) three months after THA, and the improvement rate revealed was over 70% in patients. Consequently, many improvement cases in hip joint pain were observed three months after THA. Regarding ADL-related, the improvement rate was 55% and higher in 12 questions except Q8, Q10, Q11, and Q14. These four questions mainly evaluate upper limb functions that have no substantial influence on THA. The scores before THA were 0.41 (not difficult) to 1.11 (slightly difficult), which were low to begin with, so the percentage of patients who improved their scores tended to be low. All other questions in this category evaluate locomotion movements. There were many improvements observed due to THA and rehabilitation. Regarding questions related to mental health, both Q24 (anxiety about falling) and Q25 (anxiety about difficulty in walking) were less than one point (slightly anxious) after three months of THA, indicating significant improvement. Q21 (sports activities) is one of the questions regarding social functions. It indicates the most active mobility of ADL, which is the most difficult part for hip OA patients. The score for Q21 was 2.93, at the level of “quite difficult” before THA, the highest of all questions. Three months after THA, the score decreased to 1.70, indicating a high improvement rate of 70.7%. The above results suggest that pain reduction and improved mobility have alleviated the anxiety of falling and difficulty in walking, which has led to improvement in sports activities.

Seichi et al. [15] reported that the questions of the GLFS-25 reflect the existence of multidimensional and interrelated structures in clusters. According to their study, based on seven questions (Q12, 13, 15, 16, 17, 19, 20), one cluster was formed consisting of questions associated with physical functions of ADL (walking and housework activities). This cluster presents significant relationships with other clusters and is demonstrated to be a key domain. The results from our present study revealed that the improvement rate for all seven questions was 55% and higher. Among them, the improvement rate for questions related to walking and stair climbing (Q12, 13, 15, 16, 17) ranged from 60.8% to 80.2%. For questions related to household activities (Q19, 20), the improvement rate remained at 57.9% and 60.1%. These results indicate that, among motor functions, locomotion movements showed improvements ahead of housework activities three months after THA.

The present study has two limitations that need to be considered. Firstly, both unilateral and bilateral hip OA were included in the study. Secondly, both AMIS and transgluteal surgical procedures were included. Therefore, in our further research, it is necessary to investigate whether these factors may have an impact on the improvement effect of LS.

## Conclusions

In this study of 273 patients who underwent unilateral primary THA due to hip OA, 132 patients (improvement rate 48.4%) demonstrated improvement in their total CDL stage three months after THA. Additionally, 15 out of 25 in the GLFS-25 showed improvements in over 60% of the patients. Based on these results, approximately half of the patients showed improvement in LS three months after THA. These findings underscore the significant impact of THA on improving LS, particularly in improving locomotion movements. The results emphasize the pivotal role of postoperative rehabilitation, suggesting the need to set target rehabilitation goals for enhancing motor function after THA in hip OA patients.

## Data Availability

All relevant data are within the manuscript and its Supporting Information files.

## Acknowledgments

The authors would like to thank Masaru Ochiai, Tsubasa Kawaguchi, Aya Unoki, Wakaba Iha, Akari Nagatomo, Kensuke Okamura and Mami Kanno for their assistance in recruiting the participants and for useful advice.

